# Epidemiological dynamics of SARS-CoV-2 VOC Gamma in Rio de Janeiro, Brazil

**DOI:** 10.1101/2021.07.01.21259404

**Authors:** Filipe Romero Rebello Moreira, Mirela D’arc, Diana Mariani, Alice Laschuk Herlinger, Francine Bittencourt Schiffler, Átila Duque Rossi, Isabela de Carvalho Leitão, Thamiris dos Santos Miranda, Matheus Augusto Calvano Cosentino, Marcelo Calado de Paula Tôrres, Raíssa Mirella dos Santos Cunha da Costa, Cássia Cristina Alves Gonçalves, Débora Souza Faffe, Rafael Mello Galliez, Orlando da Costa Ferreira Junior, Renato Santana Aguiar, André Felipe Andrade dos Santos, Carolina Moreira Voloch, Terezinha Marta Pereira Pinto Castiñeiras, Amilcar Tanuri, on behalf of the COVID-19-UFRJ Workgroup

## Abstract

The emergence and widespread circulation of SARS-CoV-2 variants of concern (VOC) or interest (VOI) imposes an enhanced threat to global public health. In Brazil, one of the countries most severely impacted throughout the pandemic, a complex dynamics involving variants co-circulation and turnover events has been recorded with the emergence and spread of VOC Gamma in Manaus in late 2020. In this context, we present a genomic epidemiology investigation based on samples collected between December 2020 and May 2021 in the second major Brazilian metropolis, Rio de Janeiro. By sequencing 244 novel genomes through all epidemiological weeks in this period, we were able to document the introduction and rapid dissemination of VOC Gamma in the city, driving the rise of the third local epidemic wave. Molecular clock analysis indicates this variant has circulated locally since the first weeks of 2021 and only seven weeks were necessary for it to achieve a frequency above 70%, consistent with rates of growth observed in Manaus and other states. Moreover, a Bayesian phylogeographic reconstruction indicates VOC Gamma spread throughout Brazil between December 2020 and January 2021, and that it was introduced in Rio de Janeiro through at least 13 events coming from nearly all regions of the country. Comparative analysis of RT-qPCR cycle threshold (Ct) values provides further evidence that VOC Gamma induces higher viral loads (N1 target; mean reduction of Ct: 2.7, 95% CI = ±0.7). This analysis corroborates the previously proposed mechanistic basis for this variant enhanced transmissibility and distinguished epidemiological behavior. Our results document the evolution of VOC Gamma and provide independent assessment of scenarios previously studied in Manaus, therefore contributing to the better understanding of the epidemiological dynamics currently being surveyed in other Brazilian regions.

## 1 Introduction

Severe acute respiratory syndrome coronavirus 2 (SARS-CoV-2) emerged in China in late 2019 and rapidly spread around the globe, leading the World Health Organization (WHO) to declare a pandemic state on 11 March 2020 (Wu et al., 2020; WHO, 2020). In Brazil, the first report of SARS-CoV-2 infection occurred in late February 2020 in the state of São Paulo (de Jesus et al., 2020). Soon after that, multiple introductions from diverse locations and lineages were documented in the country (Candido et al., 2020). These events led to transmission chains that perpetuated and caused a heavy burden on Brazilian public health, accounting for more than 18 million cases and 500,000 deaths reported (as of 23 June 2021; available in https://covid.saude.gov.br//).

The predominance of two major lineages, B.1.1.28 and B.1.1.33, characterized the first SARS-CoV-2 epidemic wave in Brazil (Candido et al., 2020). This scenario started to change in the second semester of 2020, with the worldwide emergence of variants of concern (VOCs). VOCs and variants of interest (VOIs) bear mutations with biological significance, potentially associated with distinct epidemiological characteristics. Diverse VOCs have been characterized in different parts of the world, as Alpha (Pango lineage B.1.1.7; Nextclade 20I/V1), Beta (Pango lineage B.1.351; Nextclade 20H/V2), Gamma (Pango lineage P.1; Nextclade 20J/V3) and Delta (Pango lineage B.1.617.2; Nextclade 21A) in the United Kingdom, South Africa, Brazil and India, respectively (Cherian et al, 2021; Tegally et al., 2020; Volz et al., 2021; Faria et al., 2021; Naveca et al., 2021). Similarly, genomic surveillance studies support that VOI Zeta (Pango lineage P.2; Nexclade 20B/S.484K) (Voloch et al., 2021), and potential VOIs N.9 (Resende et al., 2021a) and N.10 (Resende et al., 2021b) have originated in different Brazilian regions.

Interestingly, these lineages share mutations in biologically relevant sites of the receptor-binding domain (RBD) of the spike protein. For instance, the substitutions E484K and N501Y are implicated in the distinct epidemiological characteristics reported for VOCs Alpha, Beta and Gamma (Faria et al., 2021; Tegally et al., 2020; Volz et al., 2021). Some evidences suggest that VOCs can be more transmissible (Faria et al., 2021; Naveca et al., 2021; Tegally et al., 2021; Volz et al., 2021), cause increased disease severity (Davies et al., 2021; Faria et al., 2021), and display reduced neutralization by antibodies elicited by previous infections or vaccines (Cele et al., 2021; Garcia-beltran et al., 2021).

Given these distinct epidemiological behaviors and the putative fitness advantage of VOCs, current evidence supports that they have rapidly risen in frequency, becoming predominant, and quickly spread to other regions (Tegally et al., 2020; Volz et al., 2021). This phenomenon has already been recorded in Brazil. The VOI Zeta, which emerged in Rio de Janeiro in mid-July 2020, rapidly became the primary local lineage and was exported to several regions of the country and abroad (Voloch et al., 2021). Likewise, the emergence of VOC Gamma in mid-November in Manaus is marked by its fast rise in frequency and a massive increase in the number of cases (Faria et al., 2021; Naveca et al., 2021). Currently, diverse surveillance efforts indicate that this lineage has become predominant in several country regions, replacing previously circulating variants (Barbosa et al., 2021; Faria et al., 2021; Franceschi et al., 2021; Moreira et al., 2021a; Naveca et al., 2021). Moreover, VOCs that originated elsewhere, like Alpha, Beta, and Delta, have already been detected in Brazil (Claro et al., 2021; Slavov et al., 2021; SES/MA, 2021), adding up to a complex epidemiological scenario (Santos et al., 2021; Souza et al., 2020).

As the circulation of these lineages has broad epidemiological implications for public health, including ongoing vaccination efforts in Brazil, we sought to determine the genetic background of the SARS-CoV-2 epidemic in the city of Rio de Janeiro between early December 2020 and early May 2021. Rio de Janeiro is the second major Brazilian metropolis, an essential hub for business, and a majorly connected city by air travel (IATA, 2020). By combining novel genomic and epidemiological data, we were able to characterize a comprehensive shift in the composition of the local SARS-CoV-2 epidemic population induced by VOC Gamma. In addition, we provide further evidence that this variant causes higher viral loads than previously circulating variants, reinforcing its enhanced transmissibility. Altogether, this study documents the complex epidemiological dynamics of SARS-CoV-2 in a major Brazilian metropolis and highlights a possible mechanism by which VOC Gamma dominates distinct epidemiological scenarios.

## 2 Methods

### 2.1 Study population and COVID-19 diagnostics

The study population was composed by convenience sampling among the RT-qPCR positive cases amidst individuals evaluated at the COVID-19 Diagnostic Center of the Federal University of Rio de Janeiro, between 1st December 2020 and 12 May 2021. Nasopharyngeal swab samples were collected from both nostrils, placed in viral transport medium (2 mL), and kept at 4°C until transportation to the laboratory. Total viral RNA from swab samples were extracted in a KingFisher Flex System^®^ (Thermofisher, USA), using the MagMax Viral/Pathogen Kit (Thermofisher, USA), according to manufacturer’s instructions. Viral RNA was detected using the SARS-CoV-2 (2019-nCoV) multiplex CDC qPCR Probe Assay (Integrated DNA Technologies, USA) targeting the SARS-CoV-2 N1 and N2 genes, and the human ribonuclease P (RNaseP) gene (endogenous control). The GoTaq^®^ Probe 1-Step RT-qPCR System (Promega, USA) was used, according to the manufacturer’s instructions. All reactions were performed in a 7500 Thermal Cycler (Applied Biosystems, USA).

The RT-qPCR result interpretation was as follows: Positive for SARS-CoV-2 when both targets (N1 and N2) amplified with Ct ≤ 37; Undetermined when only one target amplified with Ct ≤ 37, or both targets amplified with Ct between 37 and 40; Negative when one or both targets amplified with Ct > 40, or absence of amplification.

The present study was approved by the local ethics review committee from Clementino Fraga Filho University Hospital (CAAE: 30161620.0.0000.5257) and by the national ethical review board (CAAE: 30127020.0.0000.0068). All enrolled participants were over 18 years old and declared written informed consent.

### 2.2 Genome sequencing

In total, 278 RT-qPCR positive samples with Ct < 30, collected between 11 December 2020 and 5 May 2021, were selected for genome sequencing. The temporal distribution of samples comprehended two epidemic waves of SARS-CoV-2 in Rio de Janeiro city (COE/RJ, 2021). Sequencing was carried out using a widely employed protocol (Quick et al., 2017). Briefly, viral RNA was converted to cDNA using SuperScript III or IV (ThermoFisher, USA), followed by a multiplex amplification reaction with the ARTIC SARS-CoV-2 v3 Panel and the Q5 hotstart polymerase (NewEngland Biolabs, USA). After purification, amplicons for each sample were normalized and converted into Illumina sequencing libraries using either the Nextera XT library kit (Illumina, USA) or the QIAseq FX library kit (QIAGEN, Germany), following the manufacturers’ protocols. Library fragments were quantified using Qubit dsDNA High Sensitivity assay (ThermoFisher, USA) and/or the QIAseq Library Quantification kit (QIAGEN, Germany) and their lengths were estimated using Bioanalyzer High Sensitivity DNA Analysis kit (Agilent, USA). Finally, libraries were diluted into equimolar pools and sequenced in five distinct Illumina MiSeq runs with two V2 Nano (300 cycles) and three V3 (600 cycles) cartridges.

### 2.3 Viral genome assembly

Sequencing reads were filtered with fastp v0.20.1 (Chen et al., 2018), which removed adapters, short reads (< 50 nucleotides) and trimmed low-quality bases (phread < 30). Reads from each sample were mapped against the SARS-CoV-2 reference genome (NCBI accession: NC_045512.2) with Bowtie2 v2.4.2 (Langmead and Salzberg, 2012), and mapping files were indexed and sorted with SAMtools v1.12 (Li et al., 2009). BCFtools v1.12 was used for variant calling and consensus genome inference, while BEDtools v2.30.0 (Quinlan and Hall, 2010) was used to mask low coverage sites (< 100-fold). Sequences with less than 70% genome coverage were removed from downstream analysis.

### 2.4 Lineage identification and phylogenetic analysis

Lineage identification was performed with the pangolin tool v2.4.2 (Rambaut et al., 2020; O’Toole et al., 2020). To further confirm lineage assignments and contextualize the novel sequences, a representative global dataset of SARS-CoV-2 genome sequences was assembled (*n* = 3,609). This dataset is drawn from a random sample of a larger dataset (*n* = 10,838; high-quality data available on GISAID on 13 May 2021) that comprehends all Brazilian sequences, plus one international sequence per country per epidemiological week since the first reported SARS-CoV-2 genome. These sequences were all aligned to the genome sequences herein described with MAFFT v7.475 (Katoh and Standley, 2013) and a maximum likelihood tree was inferred with IQ-tree v2.0.3 (Minh et al., 2020) under the GTR+F+I+G4 model (Tavaré, 1986; Yang, 1994).

### 2.5 Phylodynamics

To further access the temporal dynamics of introduction of VOC Gamma in Rio de Janeiro city, we performed molecular clock analyses on a fully Bayesian framework using BEAST v1.10.4 (Suchard et al., 2018). To assemble the references dataset, all Brazilian VOC Gamma sequences available on GISAID were downloaded (*n* = 3,398, as of 18 May 2021) and categorized into five discrete locations, matching Brazilian geopolitical regions: Southeast (*n* = 2,090, except Rio de Janeiro), South (*n* = 107), Northeast (*n* = 106), North (*n* = 138) and Central West (*n* = 177). The 113 novel Gamma genome sequences from Rio de Janeiro characterized in this study were added to a subset of this dataset. All sequences from locations with less than 113 representatives were included, while the same number of sequences from the remaining locations were randomly sampled, composing a geographically balanced dataset (*n* = 665). Preliminary maximum likelihood phylogenetic analysis indicated 38 reference sequences of this dataset belonged to the recently identified P.1-like clades (Gräf et al., 2021) and were removed from downstream analysis. The final dataset (*n* = 627) was composed of 113, 113, 70, 105, 113 and 113 sequences from Rio de Janeiro, Southeast, South, Northeast, North and Central West, respectively.

The time scaled phylogeographic reconstruction used: *i* – the strict molecular clock model; *ii* – a uniform prior distribution (range: 8 × 10^−4^ - 10^−3^) on evolutionary rate; *iii* – the coalescent exponential growth tree prior (Laplace prior with scale 100) (Kingman, 1982); *iv* – the HKY+I+G4 nucleotide substitution model (Hasegawa et al., 1984; Yang, 1994); and *v* – a symmetric discrete phylogeographic model (Lemey et al., 2009). Eight independent chains of 50 million generations sampling every 10,000 states were performed and convergence (effective sample size > 200 for all parameters) was verified on Tracer v1.7.1 (Rambaut et al., 2018) after 10% burnin removal. Logcombiner was used to combine posterior distributions and a maximum clade credibility tree was inferred with treeannotator.

An additional set of analyses using the non-parametric coalescent skygrid tree prior (Gill et al., 2013), with 25 grid points between the date of the most recently sampled sequence (4 May 2021) and the previously estimated date of VOC Gamma emergence (15 November 2020) (Faria et al., 2021), was also performed. These grids approximately match the number of epidemiological weeks comprehended in this period. For this analysis, 6 independent runs of 100 million generations, sampling every 10,000 steps, were executed. Maximum clade credibility trees and log files are available in **Supplementary File S1**.

### 2.6 Analysis of epidemiological data

To evaluate the hypothesis that the transmissibility enhancement of VOC Gamma is due to increments in viral loads (Faria et al., 2021; Naveca et al., 2021), we performed a series of analyses based on epidemiological data (age, sex, days of symptoms at diagnosis time and Ct values) of patients infected by viral lineages identified by genome sequencing (*n* = 244 genomes). Throughout this study, the confidence level considered for hypothesis testing was α = 0.05. First, a linear regression was used to measure the association between viral lineages (VOC Gamma or non-Gamma) and Ct values (N1 target). This model was also adjusted to account for the effects of age, sex and number of symptomatic days at diagnosis time.

We also evaluated the time series of all Ct values (N1 and N2 targets, RNaseP control) characterized in the studied period (*n* = 1,224), aiming to identify variations of viral loads related to the predominating lineages. To perform a direct comparison, we selected epidemiological weeks for which sequenced samples have shown frequency of VOC Gamma below 20% or above 80%, eliminating intermediate frequency periods. Lineages were then imputed to samples according to the epidemiological week they have been collected, and a linear model was estimated to measure the effect of lineage on Ct values. These analyses have been performed with the R software (R core team, 2020).

## 3 Results

### 3.1 Diagnostics and viral genome sequencing

A total of 1,224 individuals were positive for SARS-CoV-2 detection by RT-qPCR between 1 December 2020 and 12 May 2021 at the COVID-19 Diagnostic Center of the Federal University of Rio de Janeiro. From these, 278 samples collected between 11 December 2020 and 5 May 2021 were selected for complete genome sequencing. In this effort, 244 novel genome sequences with coverage greater than 70% (median coverage: 97.2%, range: 71.1 - 99.8%; median depth: 891-fold, depth range: 168 - 14,595-fold) were characterized, which corresponds to 19.93% of all positive individuals in the studied period. The generated data accounts for 52.5% (244/465) and 14.3% (244/1,711) of all genome sequences available on GISAID EpiCoV database for the city and state of Rio de Janeiro, respectively (as of 30 May 2021). Metadata associated with samples are available in **Supplementary File S2**, including complete sequencing statistics.

### 3.2 Analysis of viral genomes reveals a VOC Gamma driven lineage replacement event

Diverse lineages were identified in the novel genome sequences with the pangolin tool: P.2 (43.44%; 106/244), P.1 (36.47%; 89/244), B.1.1 (5.73%; 14/244), B.1 (3.69%; 9/244), B.1.1.28 (3.28%; 8/244), P.1.2 (3.28%; 8/244), B.1.1.33 (1.23%; 3/244), B.1.1.7 (1.23%; 3/244), P.4 (0.82%; 2/244), B.1.407 (0.41%; 1/244) and N.9 (0.41%; 1/244) (**Supplementary File S2**). To confirm these classifications and further contextualize these data, we performed a maximum likelihood phylogenetic inference with a globally representative dataset, which showed a different proportion of identified lineages: P.2 (46.72%; 114/244), P.1 (46.31%; 113/244), B.1.1.28 (3.68%; 9/244), B.1.1.33 (1.23%; 3/244), B.1.1.7 (1.23%; 3/244) and N.9 (0.82%; 2/244) (**Figure 1A**).

**Figure 1:**
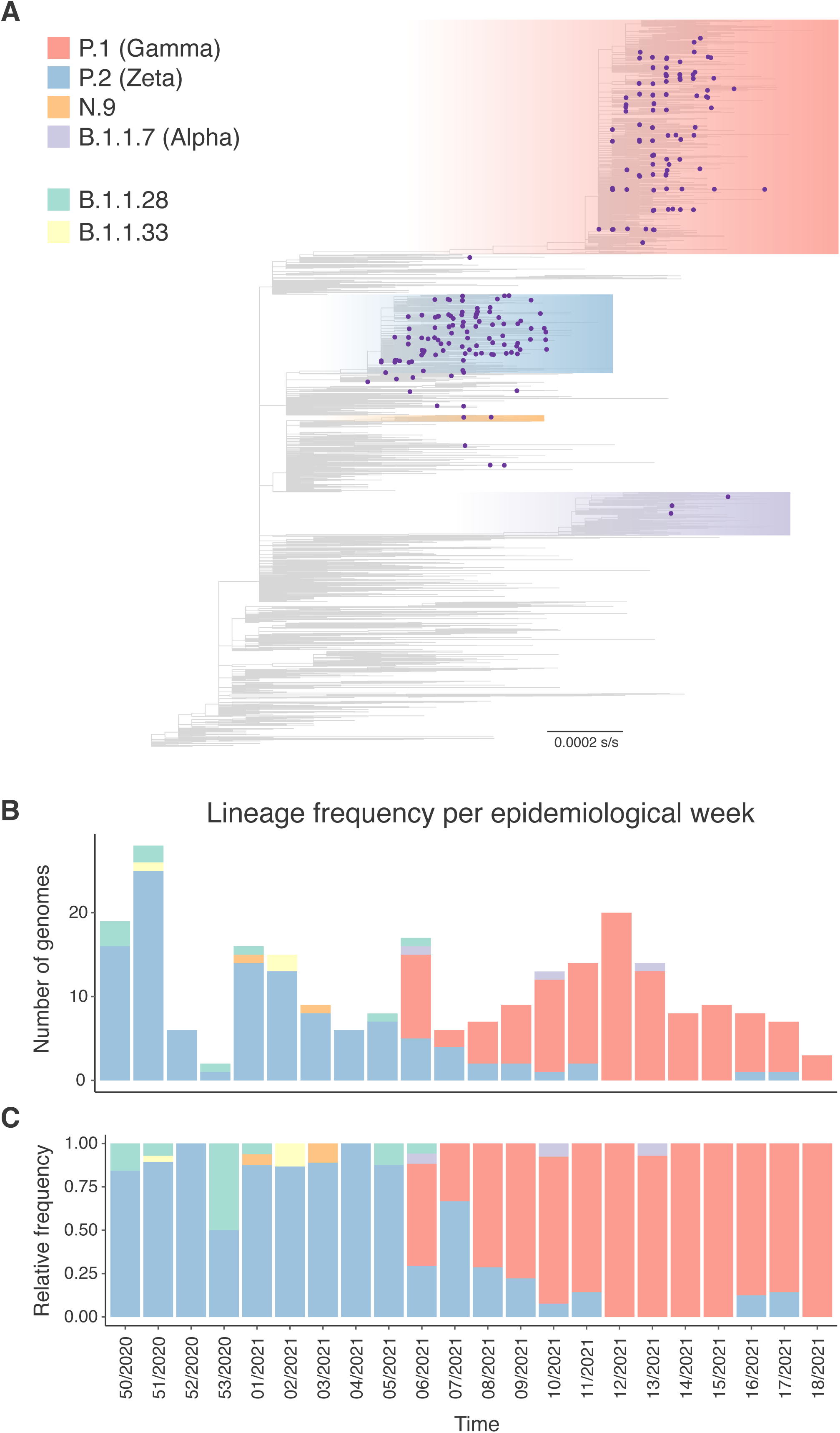
Genetic composition of SARS-CoV-2 lineages in Rio de Janeiro city in the sampled period. (A) Maximum likelihood phylogenetic tree estimated for lineage identification with a globally representative dataset. Genome sequences from this study are highlighted with purple circle tips. The lineages P.1 (Gamma), P.2 (Zeta), N.9 and B.1.1.7 (Alpha) are highlighted in red, blue, orange and light purple, respectively. Branch length scale shows 0.0002 substitutions per site (s/s). Bar plots exhibit in absolute (B) and relative (C) numbers the variation in lineages’ frequencies across epidemiological weeks from late 2020 to May 2021, as estimated from the novel SARS-CoV-2 genome sequences. Lineages are color labelled as in the phylogenetic tree, plus: B.1.1.28 (green) and B.1.1.33 (yellow).

This analysis indicates divergent classifications between the pangolin tool and the phylogenetic reconstruction, emphasizing that the latter analysis is important to confirm lineage classifications, especially for incomplete genome sequences. Sequences identified phylogenetically as P.1, P.2, B.1.1.28, and N.9 were previously misclassified as belonging to other lineages (**Supplementary File S2**).

Noticeably, the genetic composition of the SARS-CoV-2 epidemic population in Rio de Janeiro varied in the studied period (**Figures 1B and C**). The lineage P.2 (VOI Zeta) was predominant in December 2020 and January 2021, a pattern that changed with the introduction of lineage P.1 (VOC Gamma), first appearing in our dataset in the sixth epidemiological week of 2021. Since then, this lineage increased in frequency and rapidly became the major circulating lineage in the city, reaching 100% of the sampled sequences by the twelfth epidemiological week of 2021, with little variation after that. These analyses reveal a complex scenario of co-circulation of multiple VOCs or VOIs in Rio de Janeiro and highlight a fast lineage displacement event induced by VOC Gamma.

### 3.3 Phylogeographic reconstructions suggest multiple introductions of VOC Gamma into Rio de Janeiro

To further explore the temporal dynamics of introduction and spread of VOC Gamma in Rio de Janeiro city, we performed two Bayesian time scaled phylogeographic reconstructions using different demographic priors. The model based on the coalescent exponential growth prior is initially presented and the one based on the non-parametric coalescent skygrid is described below. Results from both models are summarized in **Supplementary Table S1**.

The first model places the time for the most recent common ancestor (tMRCA) of VOC Gamma on 5 November 2020 (95% highest posterior density interval, HPD: 11 October 2020 - 23 November 2020) in the North region of the country (**Figure 2A**). After emergence, the phylogeographic model suggests the variant spread to all Brazilian regions through multiple independent events occurring since late 2020. In Rio de Janeiro, 13 separate introductions that led to the emergence of local clades have been reconstructed, coming from the Northeast (*n* = 6), Central West (*n* = 2), South (*n* = 1) and Southeast (*n* = 4) regions (**Figure 2B**). A total of 7 introductions represented by single sequences have also been identified (South: 2; Southeast: 5).

**Figure 2:**
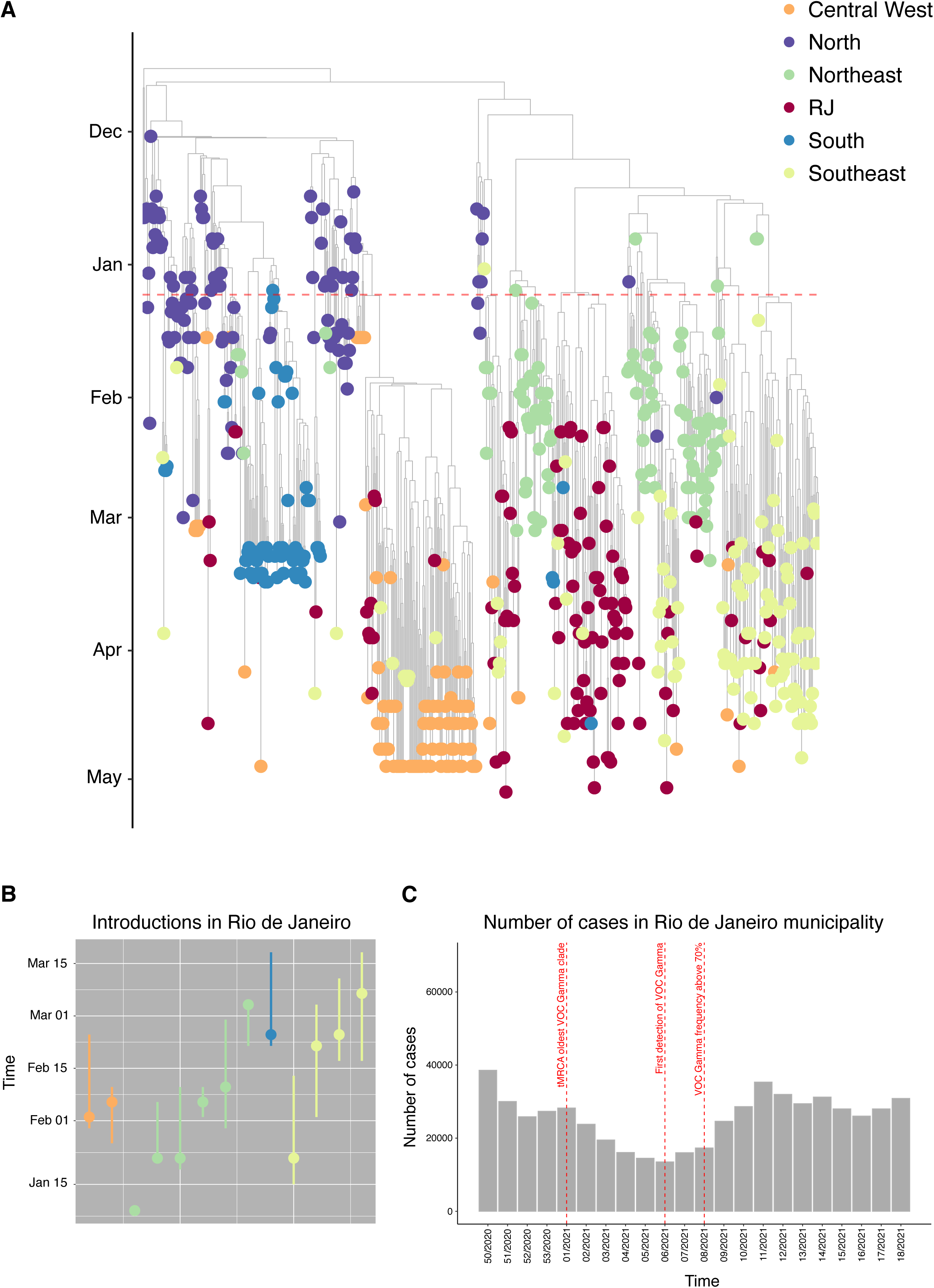
Temporal dynamics of introduction and spread of VOC Gamma in Rio de Janeiro municipality. (A) Phylogeographic reconstructions using a model based on coalescent exponential growth tree prior inferred from a dataset comprehending 514 publicly available VOC Gamma sequences and the 113 new genomes characterized in this study. (B) Dot plot showing the time of each independent VOC Gamma introduction that led to the emergence of local clades. (C) Time series of the general number of cases highlighting the VOC Gamma related events. The number of cases was obtained from the Rio COVID-19 panel (available on https://experience.arcgis.com/experience/38efc69787a346959c931568bd9e2cc4, last accessed 22 June 2021). Sampling locations are color indicated: Central West (orange), North (purple), Northeast (green), RJ (red), South (blue) and Southeast (yellow). The dashed red lines on phylogeographic reconstructions and time series highlight: 1- the time of the most recent common ancestor (tMRCA) for the oldest VOC Gamma clade (showed on both panels); 2-first detection of VOC Gamma in Rio de Janeiro (only in time series); and 3-VOC Gamma frequency above 70% (only in time series).

The model also implies that the first introduction came from the Northeast region in early January (95% HPD: 7 - 9 January 2021), after an initial dissemination of the VOC Gamma from the northern region. Posterior independent introductions from other regions of the country have been estimated over the following months (earliest event: 22 January 2021, 95% HPD: 22 January 2021 - 6 February 2021; oldest event: 7 March 2021, 95% HPD: 17 February 2021 - 18 March 2021). Likewise, exportation events from Rio de Janeiro have also been estimated, to the Central West (*n* = 2), Southeast (*n* = 9), and South (*n* = 1) regions. This molecular clock analysis provides context for the last upsurge in the number of SARS-CoV-2 infections reported in the city, mainly driven by VOC Gamma (**Figure 2C**).

The model based on the non-parametric coalescent skygrid tree prior yielded similar results, dating the emergence of VOC Gamma to 2 December 2020 (95% HPD: 23 November 2020 - 4 December 2020) in the North region. The model supports that this variant spread from the North to different regions of the country in the following months, being first introduced in Rio de Janeiro in early 2021 (**Supplementary Figure S1**). After this initial event, 13 other introductions from diverse locations have been estimated (Central West: 1; North: 4; Northeast: 2; South: 1; Southeast: 5) between January and March 2021 (earliest event: 19 January 2021, 95% HPD: 11 January 2021 - 6 February 2021; oldest event: 7 March 2021, 95% HPD: 17 February 2021 - 7 March 2021). In addition, 11 introductions supported by single sequences have also been estimated (South: 4; Southeast: 7).

The evolutionary rate estimated under the exponential model was 9.27 × 10^−4^ substitutions per site per year (s/s/y) (95% HPD: 8.53 × 10^−4^ - 9.99 × 10^−4^) and the coalescent exponential growth rate has been estimated as 14.82 (95% HPD: 12.95 - 16.76), which implicates a doubling time of 0.047 years or approximately 17 days (15 - 19 days, considering the 95% HPD). Similarly, results obtained with the coalescent skygrid tree prior indicated an evolutionary rate of 8.93 × 10^−4^ s/s/y (95% HPD: 8.10 × 10^−4^ - 9.73 × 10^−4^) and a steady growth in relative genetic diversity until early 2021, which stabilized over the following months (**Supplementary Figure S2**).

### 3.4 Analysis of RT-qPCR Ct values suggests VOC Gamma is associated with higher viral loads

To explicitly address the hypothesis that the transmissibility enhancement characterized for VOC Gamma is associated with the induction of higher viral loads, we compared the distribution of Ct values (N1 target) measured for patients infected by VOC Gamma or non-Gamma viruses. First, we conducted an exploratory data analyses to evaluate the impact of sex, age and symptomatic days at diagnosis time on Ct values using generalized linear models. While the analysis of age and days of symptoms yielded models with statistical significance, despite moderate effects, no association between patient sex and Ct values could be observed (age: *p* < 0.01, β = -0.05, 95% CI = ±0.03; days of symptoms: *p* < 0.01, β = 0.48, 95% CI = ±0.16; sex: *p* > 0.05). In this sense, multivariate linear models were performed to estimate the effect of viral lineage on Ct values, also adjusting for the individual and combined effects of age and days of symptoms. All models have been summarized in **Table 1**. No statistical association between viral lineages and Ct values could be established with statistical significance in any of the evaluated models (*p* > 0.05 for lineages, all models) (**Figure 3A**).

**Table 1:** Linear models estimated to evaluate the impact of sex, age, viral lineage and symptomatic days at diagnosis time on N1 target Ct values of sequenced samples.

| Model | Parameters | *p* - value | $\beta$ | 95% CI |
| --- | --- | --- | --- | --- |
| m1 | Days of symptoms | *<0.01* | *0.48* | *$\pm 0.16$* |
| m2 | Age | *<0.01* | *-0.05* | *$\pm 0.04$* |
| m3 | Sex | 0.14 | - | - |
| M1 | Lineage | 0.09 | - | - |
| M2 | Lineage | 0.33 | - | - |
|  | Days of symptoms | *<0.01* | *0.47* | *$\pm 0.16$* |
| M3 | Lineage | 0.07 | - | - |
|  | Age | *<0.01* | *-0.05* | *$\pm 0.03$* |
| M4 | Lineage | 0.29 | - | - |
|  | Days of symptoms | *<0.01* | *0.46* | *$\pm 0.16$* |
|  | Age | *<0.01* | *-0.05* | *$\pm 0.03$* |

Statistically significant values highlighted in italic.

**Figure 3:**
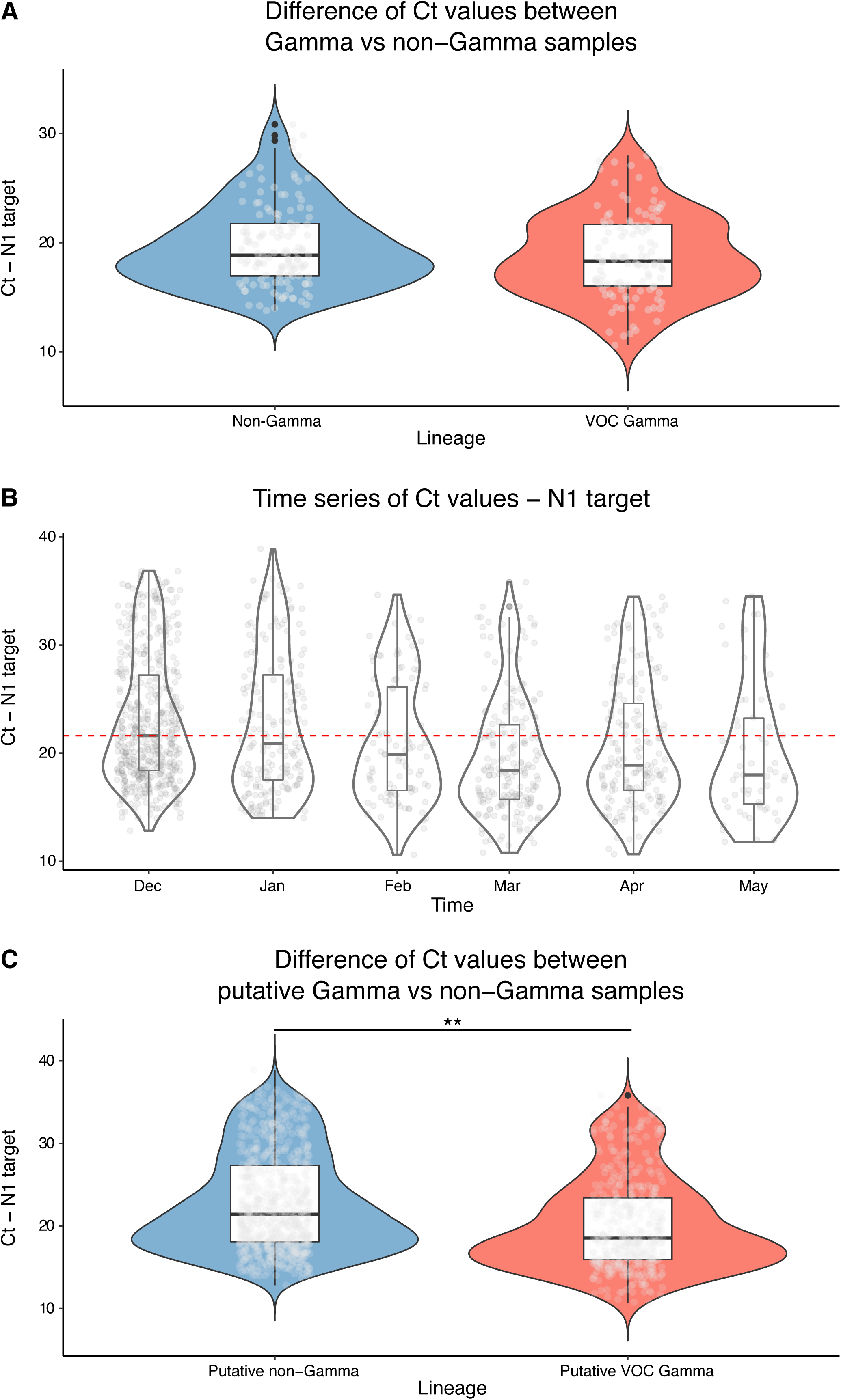
Comparison of the distribution of Ct values (N1 target) measured for patients infected by VOC Gamma or non-Gamma viruses. (A) Violin plots displaying the distribution of Ct values for different viral lineages, as characterized by genome sequencing. (B) Time series of Ct values for all positive samples in our cohort. The dashed red line represents the median Ct value observed in December 2020. (C) Violin plot displaying the distribution of Ct values for groups of samples imputed with distinct viral lineages based on collection dates. Samples collected prior to the sixth epidemiological week of 2021 were imputed as non-Gamma, while samples collected from the tenth week forward were imputed as VOC Gamma. Asterisks indicate significant statistical association between imputed viral lineages and Ct values (linear model: *p* < 0.01, β = - 2.69, 95% CI = ± 0.70).

To further examine the effect of lineages on viral loads, we evaluated the time series of Ct values (N1 target) for all positive individuals in our cohort (*n* = 1,224; **Figure 3B**). We hypothesized that if VOC Gamma in fact induces higher viral loads, a continuous decrease in Ct values should be observed as its frequency increases. This pattern is consistent with the observed global Ct values between December 2020 (median Ct value: 21.61) and March 2021 (median Ct value: 18.37), in agreement with the rise in frequency of VOC Gamma. Nevertheless, this tendency moved slightly upwards in April 2020 (median Ct value: 18.88), suggesting other factors could also be influencing Ct values distribution. Time series analysis of Ct values for the N2 target yielded similar results, while for the RNaseP control no tendencies could be identified, validating previous results (**Supplementary Figure S3**).

To perform a direct comparison, lineages have been imputed to all positive samples based on their frequency per epidemiological week, estimated with sequencing data. Samples collected before the sixth epidemiological week of 2021 (*n* = 749; median N1 target Ct value: 21.43) were imputed as non-Gamma, while samples collected from tenth epidemiological week of 2021 forward (*n* = 420; median N1 target Ct value: 18.54) were imputed as VOC Gamma. This comparison suggests the existence of a significant statistical association between lineages and Ct values (*p* < 0.01, β = -2.69, 95% CI = ±0.70) (**Figure 3C**). This result was further corroborated for the N2 target (*p* < 0.01, β = -2.43, 95% CI = ±0.75) and no significant statistical association was identified for RNaseP (*p* = 0.83).

## 4 Discussion

Brazil has been severely impacted by the SARS-CoV-2 pandemic, accounting for 9.5% of the cases and 10.4% of the deaths reported worldwide, even though the country harbors approximately 2.7% of the global population (Castro et al., 2021). While the epidemiological dynamics on a national scale has been shown to be affected by a range of complex factors (Santos et al., 2021; Souza et al., 2020), the scenario is further aggravated by the emergence and widespread circulation of multiple VOCs and VOIs in the country (Claro et al., 2021; Faria et al., 2021; Naveca et al., 2021; Resende et al., 2021a; Resende et al., 2021b; Slavov et al., 2021; Voloch et al., 2021; SES/MA, 2021). As these lineages harbor a myriad of mutations of biological significance and epidemiological implications (Cele et al., 2021; Davies et al., 2021; Faria et al., 2021; Garcia-beltran et al., 2021; Tegally et al., 2021; Volz et al., 2021), we sought to investigate their circulation dynamics in the city of Rio de Janeiro, the second largest Brazilian metropolis. By combining novel genetic and epidemiological data, we documented a fast lineage replacement event induced by VOC Gamma and provide additional evidence of its altered epidemiological characteristics.

Our study was conducted between early December 2020 and early May 2021, capturing the second and third epidemic waves in the city of Rio de Janeiro (COE/RJ, 2021). Analysis of the 244 novel viral genomes led to the identification of six circulating lineages by phylogenetic inference: P.1, P.2, B.1.1.28, B.1.1.7, B.1.1.33 and N.9 (**Figure 1A**). Temporal analysis of lineage frequencies revealed a shift in the genetic composition of SARS-CoV-2 epidemic population in Rio de Janeiro (**Figures 1B and 1C**), with lineage P.2 (VOI Zeta), most frequent between December 2020 and January 2021, being replaced by lineage P.1 (VOC Gamma), first identified in the sixth epidemiological week of 2021. In fact, VOC Gamma has rapidly risen in frequency, being responsible for over 70% of cases only two weeks later, coinciding with the third epidemic wave in Rio de Janeiro city (COE/RJ, 2021). This result reveals a complex epidemiological scenario, marked by co-circulation of multiple VOCs (Alpha and Gamma) and VOIs (Zeta and potential VOI N.9) and a fast lineage turnover event induced by VOC Gamma, consistent with previous reports on other Brazilian states (Barbosa et al., 2021; Faria et al., 2021; Franceschi et al., 2021; Moreira et al., 2021a; Naveca et al., 2021). Moreover, these results corroborate previous data showing an early detection of VOC Gamma in Rio de Janeiro and provide further context for preliminary genomic analysis performed for the whole state (Almeida et al., 2021; Lamarca et al., 2021).

Beyond the reported lineage displacement event, the dissemination of VOC Gamma in Rio de Janeiro is also marked by a small interval between autochthonous circulation and an associated epidemic wave. While VOI Zeta emerged in mid-July 2020 (Voloch et al., 2021) and rose in frequency through the second semester, causing an epidemic peak over the last months of that year, our molecular clock analysis indicates that only 10 weeks separate the tMRCA of the earliest Gamma clade from Rio de Janeiro and the epidemic peak of the third wave, mostly driven by this lineage (**Figures 2A and 2C**). This observation implies that VOC Gamma not only displayed higher transmissibility than VOI Zeta, but also has been associated with an upsurge in the number of cases, in close agreement with reports from Manaus (Faria et al., 2021; Naveca et al., 2021).

Except for B.1.1.28 and B.1.1.33, the predominant lineages in the first epidemic wave in Brazil (Candido et al., 2020), all the remaining lineages are VOCs, VOIs or potential VOIs and share the mutation E484K on the RBD of the spike protein, shown to weaken neutralizing antibodies response (Greaney et al., 2021). This observation highlights the previously reported evolutionary convergence on SARS-CoV-2 genomes (Martin et al., 2021). Likewise, both VOCs Alpha and Gamma share the mutation N501Y, also implicated in immune escape and enhancement of the affinity between the RBD and human angiotensin converting enzyme 2 (ACE2), the cellular receptor used by SARS-CoV-2 for cell entry (Lan et al., 2020; Ramanathan et al., 2021). These functional effects, joint to the fast observed rise in frequency and number of cases across diverse epidemiological settings (Barbosa et al., 2021; Faria et al., 2021; Franceschi et al., 2021; Moreira et al., 2021a; Naveca et al., 2021) - including the one herein reported - provide evidence that the VOC Gamma has a fitness advantage over previously circulating lineages.

These results are consistent with the singular epidemiological characteristics reported for other VOCs in distinct epidemiological scenarios, as Alpha (Volz et al., 2021) and Beta (Tegally et al., 2021). Interestingly, although both lineages have been shown to circulate in Brazil (Claro et al., 2021; Slavov et al., 2021), with evidence that VOC Alpha circulates in nearly all regions of the country (Moreira et al., 2021b), no rise in frequency for the later variant could be observed in our dataset. This result suggests these lineages do not have a fitness advantage in a scenario dominated by VOC Gamma, a hypothesis that must be evaluated across other settings.

Molecular clock analysis performed with the coalescent exponential growth and skygrid tree priors yielded only marginally different results, so the first model will be considered for the purpose of this discussion. The time scaled phylogenetic reconstruction suggests VOC Gamma emerged in the North region between middle October and late November 2020, consistent with previous reports from Manaus (Faria et al., 2021; Naveca et al., 2021). The phylogeographic model suggests that after initial dissemination to other regions of the country in early 2021, this lineage was introduced in Rio de Janeiro through multiple events from diverse locations (**Figure 2**). While the first estimated introduction came from the Northeast region in early January (95% HPD: 7 - 9 January 2021), later events from nearly all other regions in the following months have been estimated. This phylogeographic reconstruction emphasizes the mixture of viruses from diverse regions of the country, evidencing the necessity of coordinated surveillance efforts on a national scale. Although the estimated number of introduction events is certainly underestimated, given the limited number of genomes analyzed, this phylogeographic reconstruction likely reveals general patterns of viral spread across the country and, ultimately, the dynamics of VOC Gamma dissemination into Rio de Janeiro.

The molecular clock analysis also implies that it took approximately five weeks for VOC Gamma to be detected by genome sequencing in our randomly selected set of samples and seven weeks to achieve 70% frequency, consistent with previous rates of growth observed in Manaus (Faria et al., 2021). Moreover, demographic analysis based on the coalescent exponential growth model implies that this lineage had a doubling time of approximately 17 days since its emergence (**Supplementary Figure S2A**). This result is consistent with the reported epidemic growth of VOC Gamma in Brazil and provides independent evidence of this phenomenon based solely on genetic data. An equivalent growth was estimated under the non-parametric coalescent skygrid tree prior (**Supplementary Figure S2B**).

Although current evidence supports that VOC Gamma in fact has a fitness advantage over other lineages circulating in Brazil, the mechanistic basis of this phenomenon has not been unequivocally established. The main hypothesis in this regard implies this variant is more transmissible due to the induction of infections characterized by higher viral loads (Faria et al., 2021; Naveca et al., 2021), as already observed for other VOCs (Tegally et al., 2021; Volz et al., 2021). Our results agree with this conjecture. No effect of viral lineages on N1 target Ct values distributions could be determined when considering only sequenced samples (**Figure 3A**). However, the total number of samples and possible biases associated with sample selection criteria for genome sequencing (Ct < 30) limit the scope of this analysis. Notwithstanding, analysis of N1 target Ct values time series were suggestive of a consistent decrement coincident with the rise in frequency of VOC Gamma (**Figure 3B**). This result is further corroborated by the analysis of Ct values for the N2 target and the RNaseP control (**Supplementary Figure S3**).

These observations were further confirmed by analysis based on imputed viral lineages through sample collection dates, supporting that samples from individuals infected with VOC Gamma viruses have lower Ct values, *i*.*e*., higher viral loads (**Figure 3C**). These results are in line with previous estimates (Faria et al., 2021; Naveca et al., 2021) and suggest that alternative methods of lineage identification (e.g., RT-qPCR) (Naveca et al., 2021; Vogels et al., 2021) may be especially useful in detecting the effect of lineages on Ct values in an unbiased fashion.

It has been suggested that Ct values also variate as a function of epidemiological trajectories (Hay et al., 2021). To ascertain that this factor has not imposed a significant bias on our analysis, we subsetted epidemiological weeks from periods of decay of the second (40/2020 to 01/2021) and third (11/2021 to 16/2021) epidemic waves in Rio de Janeiro. Samples from the weeks of decay of the second and third waves were imputed as non-Gamma and VOC Gamma, respectively. This analysis yielded the same qualitative results (N1 target; *p* < 0.01; β = -2.96; 95% CI = ±0.81). The presented findings would benefit from further studies over broader time scales and across different epidemiological settings.

Additionally, it is conceivable that other mechanisms might also play a role in the enhancement of VOC Gamma transmissibility. For instance, assays measuring binding between ACE2 and the RBD of VOCs Alpha and Beta showed 1.98- and 4.62-fold increments on affinity in comparison to the original Wuhan isolate, respectively (Ramanathan et al., 2021). As VOC Gamma harbors multiple RBD mutations in common with VOC Beta – K417N/T, E484K, N501Y – it is likely that it also exhibits differences in the affinity with the human ACE2, enhancing its transmission potential. Future functional studies should evaluate this conjecture.

Altogether, this study describes a complex epidemiological dynamics for SARS-CoV-2 in a major metropolis of Brazil, one of the countries most severely hit by the COVID-19 pandemic (Castro et al., 2021). We document the joint circulation of multiple VOCs and VOIs and report a fast lineage displacement event induced by VOC Gamma, providing further evidence of its altered epidemiological characteristics. In addition, through evolutionary analyses, we describe the timing and origins of VOC Gamma introductions in the city. Finally, we bring forth data and analyses that support the hypothesis that the transmissibility enhancement associated with this lineage is at least partially explained by the induction of higher viral loads. Overall, our results document the evolution of this variant and provide independent assessment of scenarios previously studied in Manaus, therefore contributing to the better understanding of the epidemiological dynamics currently being observed in other Brazilian regions.

## Data Availability

All consensus genome sequences characterized in this study have been deposited on GISAID (Epicodes: EPI_ISL_2629675 to 2629820). Associated metadata are available in Supplementary File S2.

## Data availability

All consensus genome sequences characterized in this study have been deposited on GISAID (Epicodes: EPI_ISL_2629675 to 2629820). Associated metadata are available in **Supplementary File S2**.

## Acknowledgements

We would like to thank all authors who submitted their data to GISAID and the EpiCoV curation team for their work. A complete list of acknowledgements is available on **Supplementary File S3**. We would also like to thank the entire technical and administrative staff from Laboratório de Virologia Molecular for their invaluable support, making this and other studies by our group feasible. Further, we would like to acknowledge Mr. Rafael Estima and Druid for their help on handling computational resources that made our analyses possible. Finally, we would also like to thank the Instituto Nacional do Cancer staff for the technical support on part of our sequencing runs.

## Members of the COVID-19-UFRJ Workgroup

Anna Carla Pinto Castiñeiras, Bianca Ortiz da Silva, Cintia Policarpo, Cynthia Chester Cardoso, Érica Ramos dos Santos Nascimento, Fernanda Leitão dos Santos, Gleidson da Silva de Oliveira, Guilherme Sant’Anna de Lira, Helena D’Anunciação de Oliveira, Helena Toledo Scheid, Isabela Labarba Carvalho de Almeida, Laura Zalcberg Renault, Lidia Theodoro Boullosa, Marcelo Amaral de Souza, Mariana Campos Freire, Natacha Cunha de Araujo Faria, Raquel Fernandes Coelho, Ricardo José Barbosa Salviano, Romina Carvalho Ferreira, Thais Felix, Victor Akira Ota, Victoria Cortes Bastos.

## Supplementary Material

**Supplementary Figure S1:** Phylogeographic reconstructions using a model based on non-parametric coalescent skygrid tree prior. Inner panel on the right showing introductions that led to the emergence of local VOC Gamma clades in Rio de Janeiro.

**Supplementary Figure S2:** Demographic analysis based on the coalescent exponential growth model and non-parametric coalescent skygrid model. Both models imply an initial increase in relative genetic diversity, matching the reported rise in frequency of lineage P.1 across the country.

**Supplementary Figure S3:** Time series of Ct values for all positive samples in our cohort. Violin plot displaying the distribution of Ct values for N1 (A) and N2 (B) viral targets and RNaseP endogenous control (C). The dashed red line represents the median Ct value observed in December 2020.

**Supplementary Table S1:** Results from BEAST models based on the coalescent exponential growth and non-parametric coalescent skygrid tree priors.

**Supplementary File S1:** BEAST maximum clade credibility trees and log files.

**Supplementary File S2:** Metadata and associated sequencing statistics for samples characterized in this study.

**Supplementary File S3:** GISAID acknowledgement table.

