## Supplementary figures and images for "Epidemiological dynamics of SARS-CoV-2 VOC Gamma in Rio de Janeiro, Brazil"

### Supplementary Figure S1

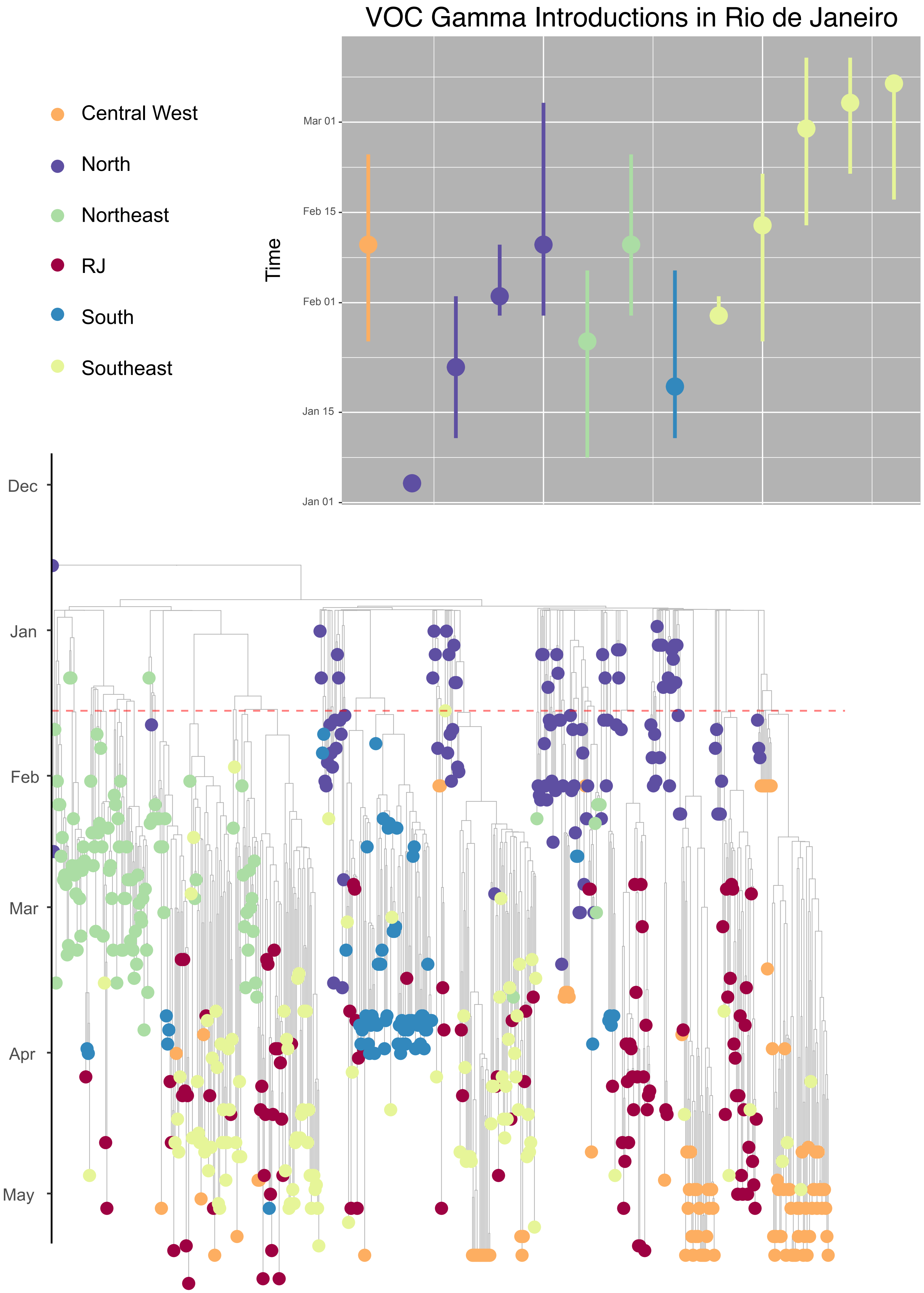

### Supplementary Figure S2

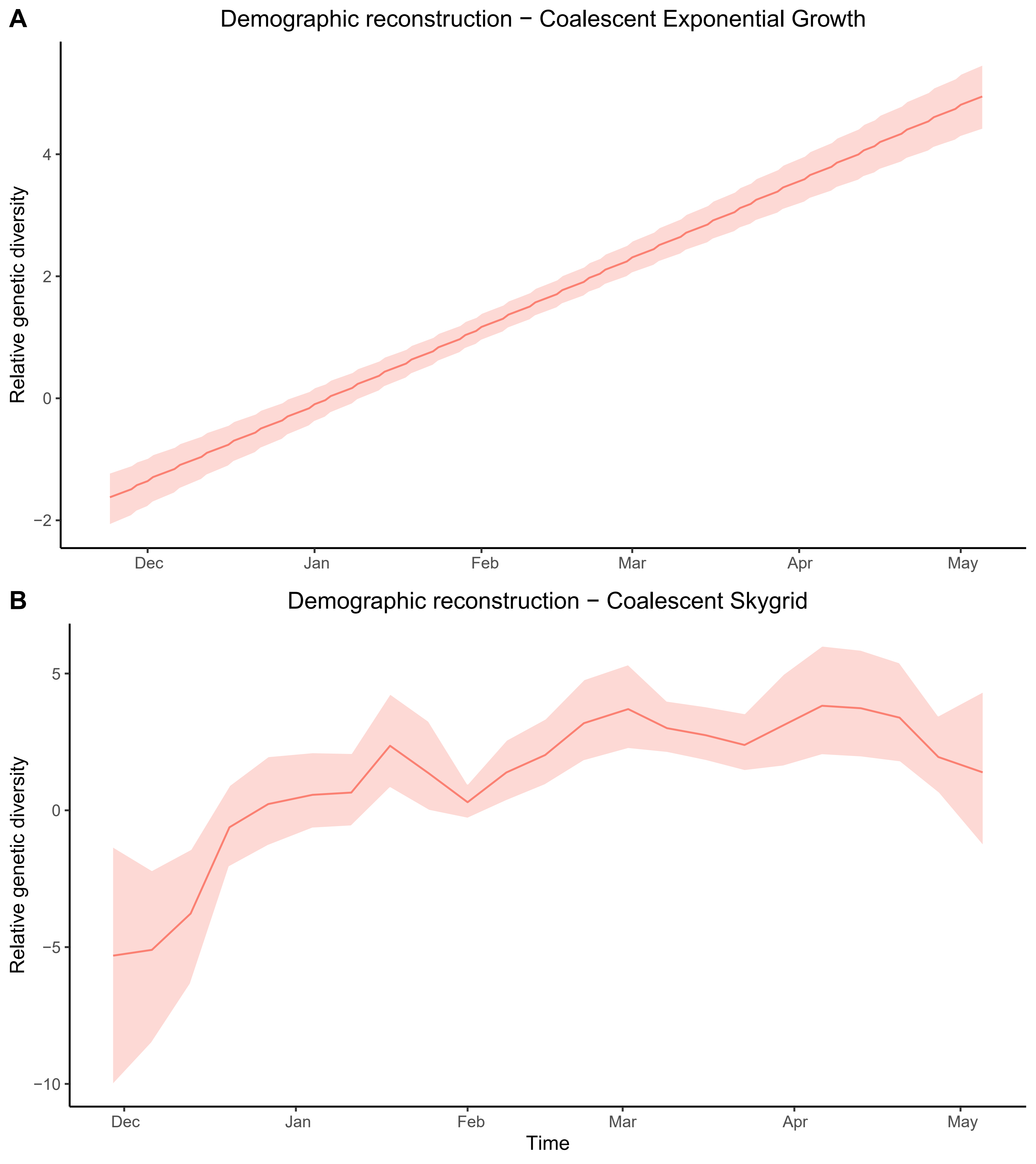

### Supplementary Figure S3

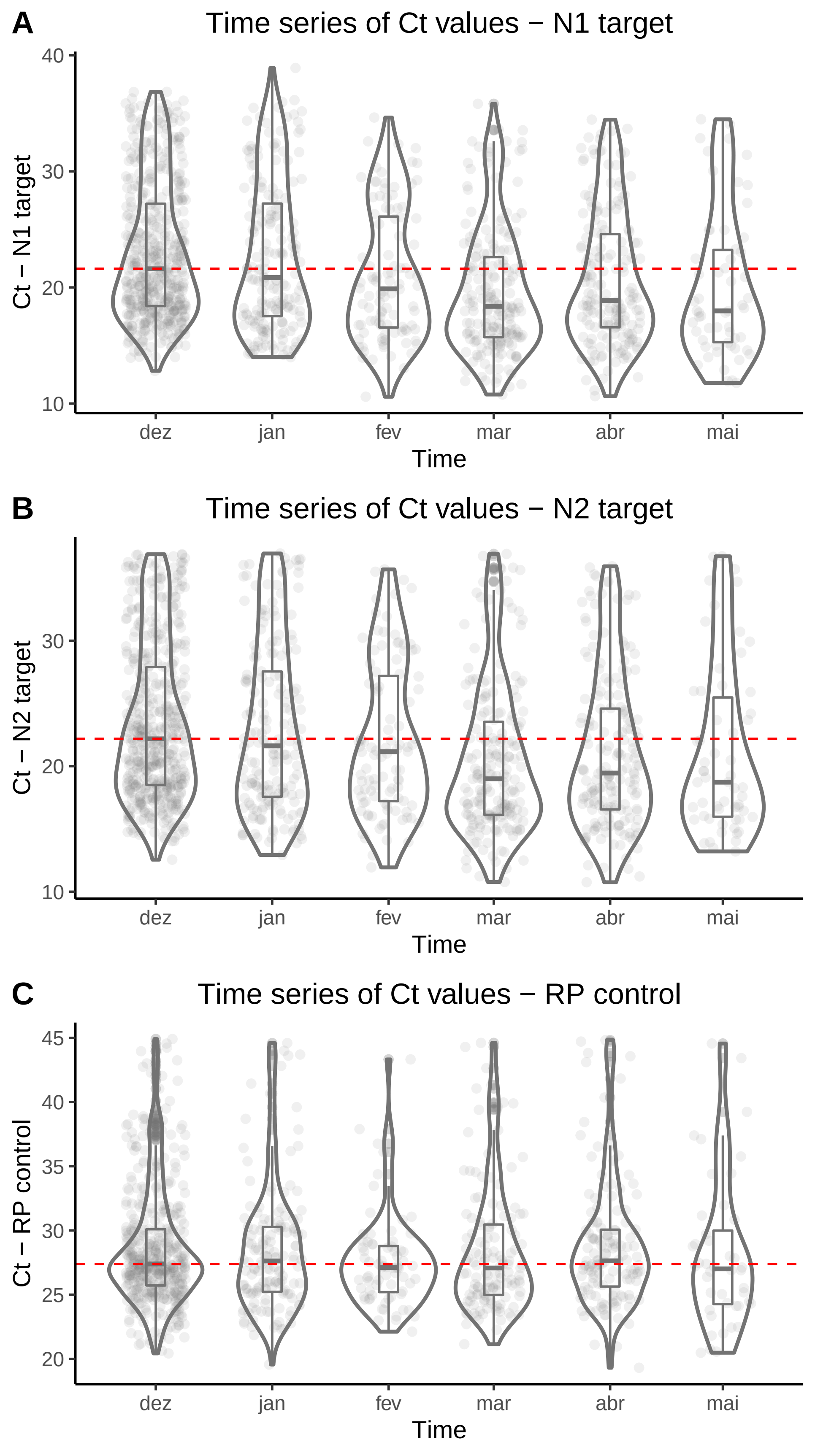
