## Supplementary Table S1 for "Epidemiological dynamics of SARS-CoV-2 VOC Gamma in Rio de Janeiro, Brazil"

**Supplementary Table S1:** Results from BEAST models based on the coalescent exponential growth and non-parametric coalescent skygrid tree priors.

| | Exponential | | | | Skygrid | | | |
|---|---|---|---|---|---|---|---|---|
| **Clade** | **tMRCA median** | **Lower boundary 95% HPD** | **Upper boundary 95% HPD** | **Location** | **tMRCA median** | **Lower boundary 95% HPD** | **Upper boundary 95% HPD** | **Location** |
| clade1 | 2021.15 | 2021.13 | 2021.19 | Southeast | 2021.01 | 2021.01 | 2021.01 | North |
| clade2 | 2021.06 | 2021.04 | 2021.12 | Southeast | 2021.11 | 2021.08 | 2021.17 | North |
| clade3 | 2021.14 | 2021.09 | 2021.17 | Southeast | 2021.06 | 2021.03 | 2021.09 | North |
| clade4 | 2021.17 | 2021.14 | 2021.17 | Northeast | 2021.09 | 2021.08 | 2021.11 | North |
| clade5 | 2021.18 | 2021.13 | 2021.21 | Southeast | 2021.18 | 2021.13 | 2021.18 | Southeast |
| clade6 | 2021.11 | 2021.08 | 2021.16 | Northeast | 2021.17 | 2021.14 | 2021.19 | Southeast |
| clade7 | 2021.02 | 2021.02 | 2021.02 | Northeast | 2021.12 | 2021.07 | 2021.14 | Southeast |
| clade8 | 2021.15 | 2021.14 | 2021.21 | South | 2021.16 | 2021.12 | 2021.19 | Southeast |
| clade9 | 2021.06 | 2021.06 | 2021.1 | Northeast | 2021.11 | 2021.07 | 2021.15 | Central West |
| clade10 | 2021.06 | 2021.05 | 2021.11 | Northeast | 2021.05 | 2021.03 | 2021.10 | South |
| clade11 | 2021.10 | 2021.07 | 2021.11 | Central West | 2021.08 | 2021.08 | 2021.09 | Southeast |
| clade12 | 2021.10 | 2021.09 | 2021.11 | Northeast | 2021.07 | 2021.02 | 2021.10 | Northeast |
| clade13 | 2021.09 | 2021.08 | 2021.15 | Central West | 2021.11 | 2021.08 | 2021.15 | Northeast |

tMRCA = time for the Most Recent Common Ancestor
HPD = Highest Posterior Density.
